# Changing Dynamics of COVID-19 in the U.S. with the Emergence of the Delta Variant: Projections of the COVID-19 Simulator

**DOI:** 10.1101/2021.08.11.21261845

**Authors:** Jagpreet Chhatwal, Yingying Xiao, Peter Mueller, Madeline Adee, Ozden O Dalgic, Mary Ann Ladd, Turgay Ayer, Benjamin P Linas

**Affiliations:** Massachusetts General Hospital Institute for Technology Assessment, Boston, MA; Harvard Medical School, Boston, MA; H. Milton Stewart School of Industrial and Systems Engineering, Georgia Institute of Technology, Atlanta, GA; Value Analytics Labs, Boston, MA; Boston Medical Center, Boston University School of Medicine

## Abstract

With the recent emergence of the B.1.617.2 (Delta) variant of SARS-CoV-2 in the U.S., many states are seeing rising cases and hospitalizations after a period of steady decline. As We used the *<u>COVID-19 Simulator</u>*, an interactive online tool that utilizes a validated mathematical model, to simulate the trajectory of COVID-19 at the state level in the U.S. COVID-19 Simulator’s forecasts are updated weekly and included in the Centers for Disease Control and Prevention (CDC) ensemble model. We employed our model to analyze scenarios where the Delta variant becomes dominant in every state.

The combination of high transmissibility of the Delta variant, low vaccination coverage in several regions, and more relaxed attitude towards social distancing is expected to result in as surge in COVID-19 deaths in at least 40 states. In several states – including Idaho, Maine, Montana, Nebraska, North Carolina, Oregon, Puerto Rico, Washington, and West Virginia – the projected daily deaths in 2021 could exceed the prior peak daily deaths under current social distancing behavior and vaccination rate. The number of COVID-19 deaths across the U.S. could exceed 1600 per day.

Between August 1, 2021, and December 31, 2021, there could be additional 157,000 COVID-19 deaths across the U.S. Of note, our model projected approximately 20,700 COVID-19 deaths in Texas, 16,000 in California, 12,400 in Florida, 12,000 in North Carolina, and 9,300 in Georgia during this period. In contrast, the projected number of COVID-19 deaths would remain below 200 in New Jersey, Massachusetts, Connecticut, Vermont, and Rhode Island.

We project COVID-19 deaths based on the current vaccination rates and social distancing behavior. Our hope is that the findings of this report serve a warning sign and people revert to wearing masks and maintain social distancing to reduce COVID-19 associated deaths in the U.S. Our projections are updated weekly by incorporating vaccination rates and social distancing measures in each state; the latest results can be found at the *<u>COVID-19 Simulator</u>* website.

## BACKGROUND

The first confirmed case of community-transmitted severe acute respiratory syndrome coronavirus 2 (SARS-CoV-2) in the United States (U.S.) was reported in February 2020.^1^ On March 11, 2020, the World Health Organization (WHO) declared the coronavirus disease 2019 (COVID-19) caused by SARS-CoV-2 a global pandemic.^2^ As of July 31, 2021, 35 million Americans have been diagnosed with COVID-19, with over 600,00 deaths attributed to the virus.^3^

In 2020, the Food and Drug Administration (FDA) authorized three vaccines against SARS-CoV-2—BNT162b2 mRNA (Pfizer-BioNTech), mRNA-1273 (Moderna), and Ad26.COV2.S (Janssen)—for emergency use in the U.S.^4-6^ As of July 31, 2021, 46.3% of Americans have received at least one vaccine dose, with 38.7% of the population fully vaccinated (as part of a two-dose regimen). However, there is considerable heterogeneity across states with respect to vaccination coverage; 27 states have a population vaccination rate under 50%. Studies have suggested that 75–90% population-level immunity is required for communities to return to pre-pandemic levels of social mixing without triggering a rise in infections.^7,8^

Due to mass vaccination, COVID-19 cases and deaths decreased in all states from March onwards. However, with the recent emergence of the B.1.617.2 (Delta) variant of SARS-CoV-2, many states are now seeing rising cases and hospitalizations. As of July 31, 2021, 82% of daily incident cases in the U.S. were caused by the B.1.617.2 variant. Several studies have estimated the basic reproduction number of the Delta variant to be between 5 and 8. ^13^ Because of differences in population-level immunity from previous infection and vaccination, as well as attitudes towards social distancing (e.g., mask use) across states, the Delta variant could disproportionately impact some states by causing a resurgence in cases and hospitalizations. Our objective was to project COVID-19 hospitalizations and deaths in each state until December 31, 2021.

## METHODS

### Model Overview

The *COVID-19 Simulator* uses a mathematical model to simulate the epidemiology of COVID-19 at the state-level in the U.S. Since May 2020, the Centers for Disease Control and Prevention (CDC) has incorporated our model outputs in its weekly COVID-19 forecasts.^14^ Input data includes reported cases and deaths, hospitalizations and ICU occupancy, vaccine administration history, and estimates for epidemiological parameters from clinical studies. The model is calibrated to reproduce historical trends in daily reported cases and deaths, and is updated weekly as new data and evidence arise. Model programming is performed in R using numerical solvers from the package ‘deSolve’.^15^

### Model Structure

The *COVID-19 Simulator* is a compartmental SEIR model with compartments for Susceptible, Exposed, Infectious, Recovered, and Dead individuals, stratified by vaccination status (unvaccinated, partially vaccinated, fully vaccinated). A schematic of the model is shown in Figure 1.

**Figure 1.**
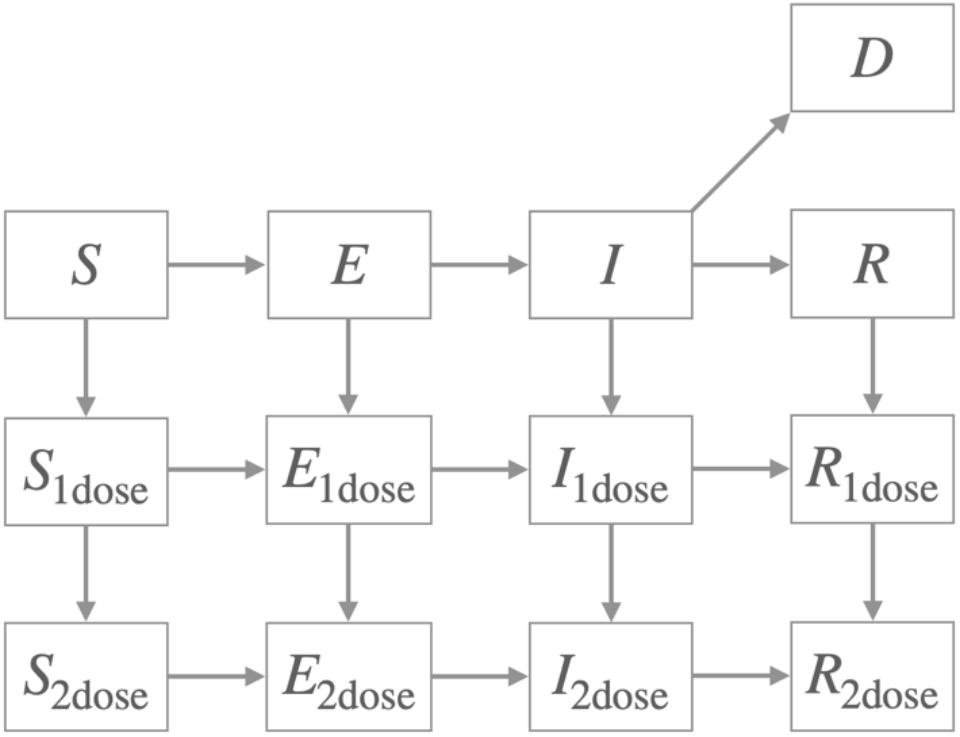
Schematic of the *COVID-19 Simulator* compartmental model. The variable subscripts ‘1dose’ and ‘2dose’ differentiate individuals in the Susceptible, Exposed, Infected, and Recovered populations who have received one or two doses of the vaccine.

### Effective Reproduction Number

The transmission force of a pandemic under a given non-pharmaceutical intervention (NPI) is described by a parameter called the *effective reproduction number, R*_*E*_, defined as the average number of secondary infections per infectious case in a population made up of both susceptible and non-susceptible people. To reproduce the historical trends in cases and deaths, we allowed *R*_*E*_ to be a function of time to capture the effect of tightening and relaxing NPIs as the pandemic progresses. Specifically, we specify a stepwise function *R*_*E*_ (*t*) with 11 joinpoints τ_1_, τ_2_, …, τ_11_ (time points at which *R*_*E*_ takes on a new value). We let the first 10 joinpoints be distributed uniformly over the historical time horizon from March 1, 2020, to the date of the latest data. We set τ_11_ to be 14 days before the date of the latest data point to capture the *R*_*E*_ of current interventions.

### Estimation of Total Infections

For all states, we use an average infection fatality rate (IFR) of 0.005 to match the estimated 115 million total infections by March 2021 estimated by the Centers for Disease Control and Prevention.^16^ The model infers the curve of total new infections (diagnosed and undiagnosed) over time based on the curve of deaths and the chosen IFR.

### Vaccine Effects

Starting from February 2021, we account for vaccine rollout. The model captures the reduced fatality rate and susceptibility of vaccinated individuals. We assume an 80% and 90% reduction in susceptibility to infection in first- and second-dose recipients respectively, along with 100% reduction in IFR for both groups.^17,18^ Due to lack of data on disease status at the time of vaccination, we conservatively assume uniform distribution of vaccines across the Susceptible, Exposed, Infected, and Recovered populations. Projecting forward, we fix vaccination rates at the latest rate from data.^19^

### Calibration of Unobserved Parameters

Because several parameters in the model are not directly observable, we estimate their values using a calibration approach. We begin by defining clinically plausible ranges:

- Initial number of infections *I*_*0*_ : 0–1,000 cases,
- Latent period duration *p*_*E*_ : 2–10 days,^20^
- Infectious period duration *p*_*1*_ : 0.1–10 days,^21^
- Effective reproduction number: 0.5–6.00 secondary cases per infectious case.

We calibrate using generalized simulated annealing from the R package ‘GenSA’ to the curve of historical new deaths as the calibration target and mean squared error as the objective function.^22^ To account for uncertainty in the calibrated values, we repeat the calibration 100 times with different random seeds, resulting in 100 unique sets of parameter values and fitted curves. At each time point, we take the median to be the point estimate and compute the 95% credible interval.

### Hospitalization and ICU Occupancy

We back-calculate hospital and ICU occupancy from incident deaths assuming an average time to death from hospital and ICU admission of 16 and 10 days respectively.^23^ Starting in August, we forecast occupancy data from HealthData.gov which notably does not cover all hospitals in a given state.^24^

### Delta Variant Scenario

For each state, we simulate the scenario where the effective reproduction number is 4.8 up to December 31, 2021.

## RESULTS

**Figure 2** shows model-estimated daily COVID-19 deaths in each state grouped by U.S. census division. The combination of high transmissibility of the Delta variant, low vaccination coverage in several regions, and more relaxed attitude towards social distancing is expected to result in a surge in COVID-19 deaths in at least 40 states.

**Figure 2.**
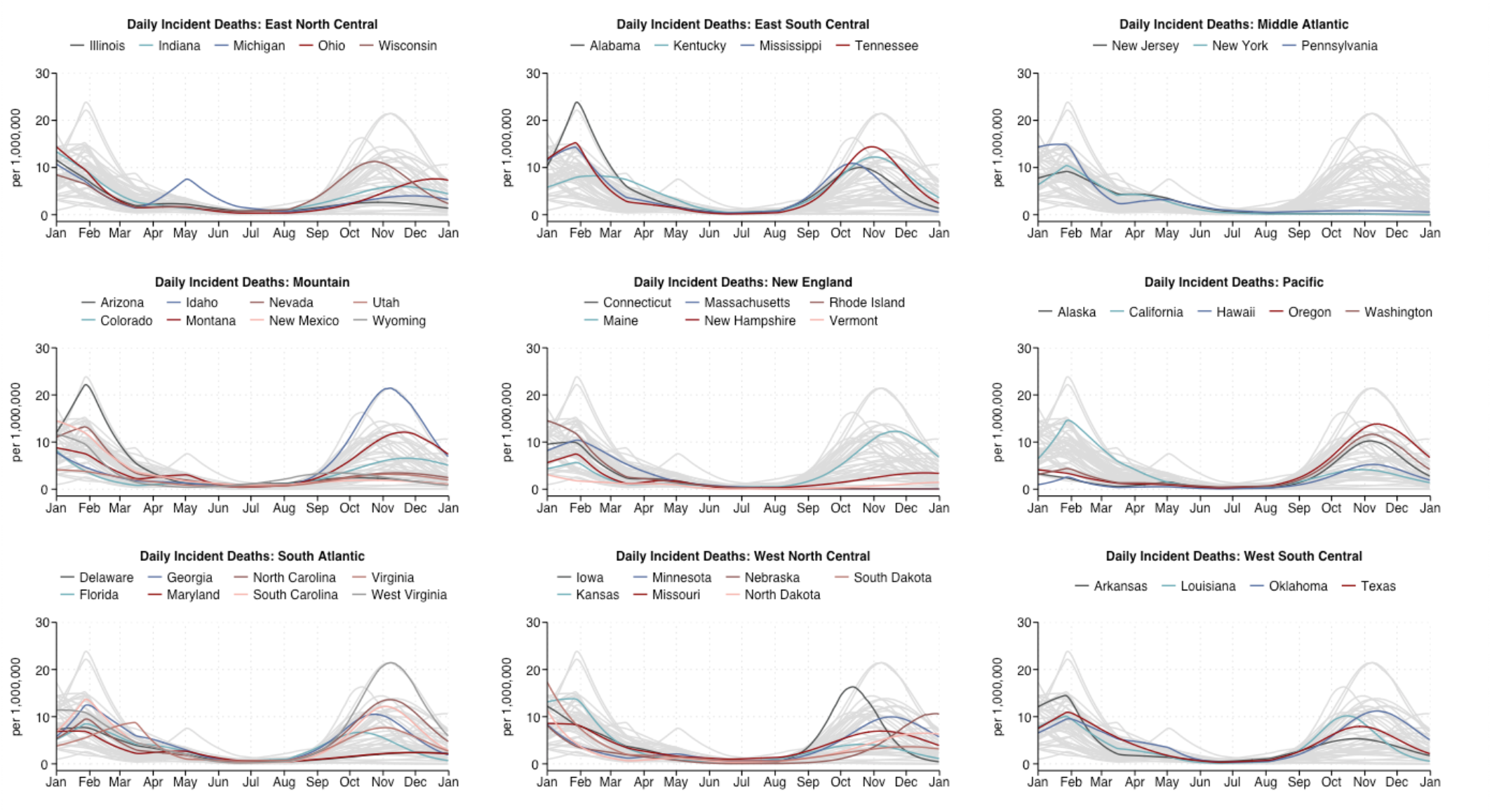
Projected COVID-19 deaths in 2021 in each state, grouped by U.S census division.

In several states––including Idaho, Maine, Montana, Nebraska, North Carolina, Oregon, Puerto Rico, Washington, and West Virginia––projected daily incident deaths could exceed their earlier 2021 peak if current social distancing behaviors and vaccination rates are maintained (**Figure 3**). Daily incident deaths across the U.S. could reach a peak of 1600 (Table 1). Of note, daily incident deaths are expected to exceed 100 in California, Florida, Georgia, North Carolina, and Texas.

**Table 1.** Projected peak and cumulative COVID-19 deaths in each state between August 1, 2021 and December 31, 2021.

|  | Cumulative Deaths | Peak Deaths |
| --- | --- | --- |
| United States | 157,089 | 1,644 |
| Alabama | 4,038 | 49 |
| Alaska | 581 | 8 |
| Arizona | 2,136 | 18 |
| Arkansas | 1,640 | 16 |
| California | 15,942 | 165 |
| Colorado | 3,812 | 38 |
| Connecticut | 63 | 0 |
| Delaware | 242 | 2 |
| District of Columbia | 37 | 0 |
| Florida | 12,358 | 143 |
| Georgia | 9,293 | 111 |
| Hawaii | 622 | 7 |
| Idaho | 3,069 | 38 |
| Illinois | 3,771 | 33 |
| Indiana | 4,287 | 40 |
| Iowa | 3,164 | 52 |
| Kansas | 1,244 | 12 |
| Kentucky | 4,710 | 55 |
| Louisiana | 3,363 | 47 |
| Maine | 1,380 | 17 |
| Maryland | 1,553 | 14 |
| Massachusetts | 124 | 1 |
| Michigan | 4,071 | 40 |
| Minnesota | 4,979 | 56 |
| Mississippi | 2,356 | 32 |
| Missouri | 4,452 | 43 |
| Montana | 1,166 | 13 |
| Nebraska | 1,092 | 21 |
| Nevada | 1,202 | 11 |
| New Hampshire | 406 | 5 |
| New Jersey | 186 | 2 |
| New Mexico | 503 | 4 |
| New York | 367 | 4 |
| North Carolina | 11,943 | 143 |
| North Dakota | 420 | 5 |
| Ohio | 6,722 | 89 |
| Oklahoma | 3,800 | 44 |
| Oregon | 5,042 | 58 |
| Pennsylvania | 1,406 | 11 |
| Puerto Rico | 1,856 | 23 |
| Rhode Island | 17 | 0 |
| South Carolina | 4,844 | 63 |
| South Dakota | 329 | 3 |
| Tennessee | 7,352 | 98 |
| Texas | 20,708 | 230 |
| Utah | 1,116 | 10 |
| Vermont | 58 | 1 |
| Virginia | 6,050 | 65 |
| Washington | 7,398 | 89 |
| West Virginia | 2,889 | 38 |
| Wisconsin | 5,708 | 66 |
| Wyoming | 219 | 2 |

**Figure 3.**
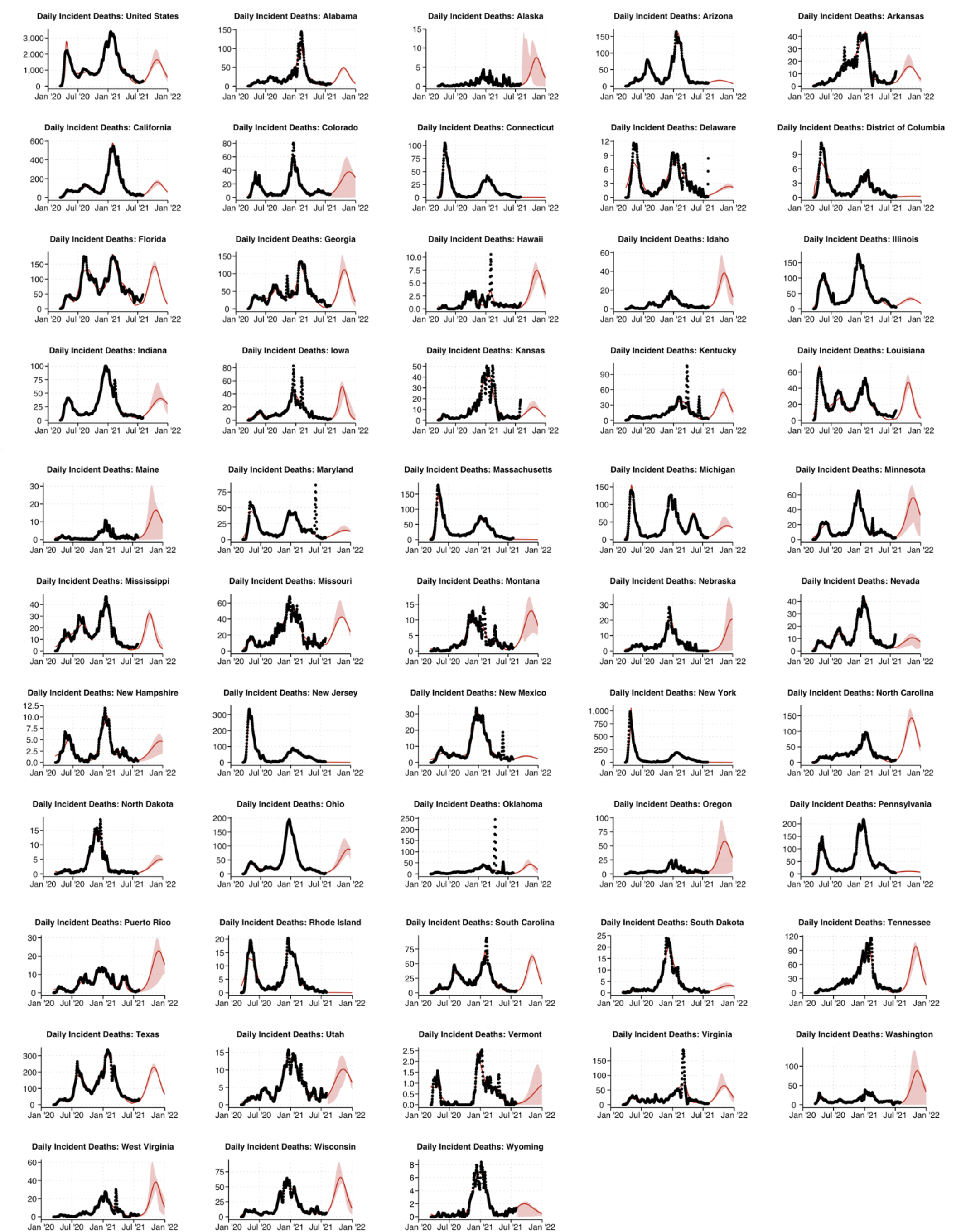
Past (black) and projected (red) COVID-19 deaths in each state until December 31, 2021.

Between August 1, 2021, and December 31, 2021, there could be an additional 157,000 COVID-19 deaths across the U.S (**Table 1**). Of note, our model forecasts 20,700 COVID-19 additional deaths in Texas, 16,000 in California, 12,400 in Florida, 12,000 in North Carolina, and 9,300 in Georgia during this period. In contrast, most states in the Northeast would not see a significant increase in deaths. The projected number of COVID-19 deaths between August 1, 2021, and December 31, 2021, is below 200 in New Jersey, Massachusetts, Connecticut, Vermont, and Rhode Island.

## Data Availability

Our model is available to the public at www.covid19sim.org and data can be downloaded from the website.

## REFERENCES

1. Jorden MA, Rudman SL, Villarino E, et al. Evidence for Limited Early Spread of COVID-19 Within the United States, January-February 2020. MMWR Morb Mortal Wkly Rep. 2020;69(22):680–684.

2. Timeline of WHO’s response to COVID-19. Retrieved from: https://www.who.int/news-room/detail/29-06-2020-covidtimeline (last accessed: Aug 5, 2020).

3. COVID Data Tracker. Centers for Disease Control and Prevention. Retrieved from: https://covid.cdc.gov/covid-data-tracker/#datatracker-home (accessed: April 8, 2021).

4. Polack FP, Thomas SJ, Kitchin N, et al. Safety and Efficacy of the BNT162b2 mRNA Covid-19 Vaccine. New England Journal of Medicine. 2020;383(27):2603–2615.

5. Baden LR, El Sahly HM, Essink B, et al. Efficacy and Safety of the mRNA-1273 SARS-CoV-2 Vaccine. New England Journal of Medicine. 2020;384(5):403–416.

6. Janssen Ad26.COV2.S Vaccine for the Prevention of COVID-19. FDA Briefing Document. Vaccines and Related Biological Products Advisory Committee Meeting. February 26, 2021. Retrieved from: https://www.fda.gov/media/146217/download (accessed: March 16, 2021).

7. Aschwanden C. The false promise of herd immunity for COVID-19. Nature. 2020;587(7832):26–28.

8. Omer SB, Yildirim I, Forman HP. Herd Immunity and Implications for SARS-CoV-2 Control. Jama. 2020;324(20):2095–2096.

9. CDC COVID-19 Forecasts. Retrieved from: https://www.cdc.gov/coronavirus/2019-ncov/covid-data/forecasting-us.html, last accessed: June 5, 2020).

10. Hansen CH, Michlmayr D, Gubbels SM, Mølbak K, Ethelberg S. Assessment of protection against reinfection with SARS-CoV-2 among 4 million PCR-tested individuals in Denmark in 2020: a population-level observational study. The Lancet. 2021.

11. Harvey RA, Rassen JA, Kabelac CA, et al. Association of SARS-CoV-2 Seropositive Antibody Test With Risk of Future Infection. JAMA Internal Medicine. 2021.

12. Lumley SF, O’Donnell D, Stoesser NE, et al. Antibody Status and Incidence of SARS-CoV-2 Infection in Health Care Workers. New England Journal of Medicine. 2020;384(6):533–540.

13. Covid: Is there a limit to how much worse variants can get? BBC News. https://www.bbc.com/news/health-57431420. Published June 11, 2021. Accessed August 10, 2021.

14. Cramer EY, Ray EL, Lopez VK, et al. Evaluation of individual and ensemble probabilistic forecasts of COVID-19 mortality in the US. medRxiv. Published online February 5, 2021:2021.02.03.21250974. doi:10.1101/2021.02.03.21250974

15. Soetaert K, Petzoldt T, Setzer RW. Package deSolve: Solving Initial Value Differential Equations in R. :52.

16. Centers for Disease Control and Prevention. Estimated Disease Burden of COVID-19. Centers for Disease Control and Prevention. Published February 11, 2020. Accessed May 15, 2021. https://www.cdc.gov/coronavirus/2019-ncov/cases-updates/burden.html

17. Thompson M, Burgess J, Naleway A, et al. Interim Estimates of Vaccine Effectiveness of BNT162b2 and mRNA-1273 COVID-19 Vaccines in Preventing SARS-CoV-2 Infection Among Health Care Personnel, First Responders, and Other Essential and Frontline Workers — Eight U.S. Locations, December 2020–March 2021. MMWR Morbidity and Mortality Weekly Report. 2021;70. doi:10.15585/mmwr.mm7013e3

18. Mahase E. Covid-19: Pfizer vaccine efficacy was 52% after first dose and 95% after second dose, paper shows. BMJ. 2020;371:m4826. doi:10.1136/bmj.m4826

19. Mathieu E, Ritchie H, Ortiz-Ospina E, et al. A global database of COVID-19 vaccinations. Nat Hum Behav. Published online May 10, 2021:1–7. doi:10.1038/s41562-021-01122-8

20. Lauer SA, Grantz KH, Bi Q, et al. The Incubation Period of Coronavirus Disease 2019 (COVID-19) From Publicly Reported Confirmed Cases: Estimation and Application. Ann Intern Med. Published online March 10, 2020. doi:10.7326/M20-0504

21. Li Q, Guan X, Wu P, et al. Early Transmission Dynamics in Wuhan, China, of Novel Coronavirus–Infected Pneumonia. New England Journal of Medicine. 2020;382(13):1199–1207. doi:10.1056/NEJMoa2001316

22. Xiang Y, Gubian S, Suomela B, Hoeng J. Generalized Simulated Annealing for Global Optimization: The GenSA Package. The R Journal. 2013;5(1):13. doi:10.32614/RJ-2013-002

23. Ferguson N, Laydon D, Nedjati Gilani G, et al. Report 9: Impact of Non-Pharmaceutical Interventions (NPIs) to Reduce COVID19 Mortality and Healthcare Demand.; 2020. doi:10.25561/77482

24. HealthData.gov. COVID-19 Reported Patient Impact and Hospital Capacity by State Timeseries | HealthData.gov. Accessed August 3, 2021. https://healthdata.gov/Hospital/COVID-19-Reported-Patient-Impact-and-Hospital-Capa/g62h-syeh

